# Evaluation of changes in coagulation factors in fresh frozen plasma during storage at -18°C for 5 weeks at Kisii Teaching and Referral Hospital

**DOI:** 10.1101/2023.04.06.23288241

**Authors:** Collince O. Ogolla, Rodgers N. Demba

## Abstract

**Background:** Fresh frozen plasma is a critical substitute therapy in management of bleeding. Increased risk of venous thrombosis has been described to be associated with high plasma levels of several coagulation factors.

**Methodology:** This study was a longitudinal study involving time series analysis of fresh frozen plasma stored at -18°C for five weeks. A sample of 180 ml plasma was obtained from the blood centrifuged at 4000rpm which was aliquoted into three parts each containing 60ml. The first aliquot was used to assess the changes in coagulation factors in FFP at baseline during the first week of sample collection, the second aliquot was used to assess the changes in coagulation factors in FFP storage at -18°C temp after three weeks of storage, the third aliquot was used to assess the changes in coagulation factors in FFP storage at -18°C temp after five weeks of storage. Coagulation factor analysis was performed using Erba Mannheim ECL 105 coagulation analyzer, India factor results recorded. Thawing for subsequent coagulation factor analysis and serial testing of stored cryoprecipitate and fresh frozen plasma was done using Stericox Plasma Thawing Bath at 37°C, for 45 mins before before analyzing the samples. Standard storage conditions for the aliquots were monitored and maintained to ensure homogeneity.

**Results:** The findings showed significant changes in the coagulation factors in FFP during storage at -18 for a period to five weeks with chi-square value of 216.000 and asymptomatic significance value (p-value) <0.0001* less than the standard alpha 0.05.

**Conclusion:** There was a constant decrease of coagulation factors in fresh frozen plasma during storage at -18°C for 5 weeks at Kisii Teaching and Referral Hospital, Kisii County.

## Introductiom

Fresh frozen plasma is the fluid portion of a unit of whole blood centrifuged, separated, and frozen solid at minus 18 °C (0 °F) or colder within eight hours of collection from whole blood donation or via apheresis device (1). Fresh frozen plasma is stored at -18ºC or below and thawed in a water bath at 30 to 37ºC for 20 to 30 minutes or in an FDA-cleared device as quickly as 2 to 3 minutes before being administered and administered immediately after thawing. Thawed fresh frozen plasma is stored at 1 to 6ºC if not given immediately after thawing and discarded if not used in 24 hours. Coagulation factors are proteins withinside the blood that assist to manage bleeding (1). The factors are generally enzymes (serine proteases) which act by cleaving downstream proteins. Coagulation system is a highly regulated cascade which involves intrinsic and extrinsic pathways leading to blood clot formation (1). The clotting factors includes Factor I (fibrinogen), Factor II (prothrombin), Factor III (tissue thromboplastin or tissue factor), Factor IV (ionized calcium), Factor V (labile factor or proaccelerin), Factor VII (stable factor or proconvertin), and Factor VIII (antihemophilic factor), Factor IX (plasma thromboplastin component or the Christmas factor), Factor X (Stuart-Prower factor), Factor XI (plasma thromboplastin antecedent), Factor XII (Hageman factor), and Factor XIII (fibrin-stabilizing factor) (2). Several physiological and pathological variables affect coagulation factor levels (3).

The activity of clotting factors, predominantly factor VIII and factor V, decline gradually once thawed (4). Fresh Frozen Plasma has specific indications which are limited to the treatment of deficiencies of coagulation proteins for which specific factor concentrates are undesirable or unavailable (5). FFP is indicated in the replacement of isolated factor deficiencies and reversal of warfarin effect, treatment of thrombotic thrombocytopenic purpura, for antithrombin III deficiency, treatment of immunodeficiencies, treatment of conditions in which there are low blood clotting factors or low levels of other blood proteins and replacement fluid in plasma exchange. Fresh frozen plasma contains all of the clotting factors (5).

Changes in plasma coagulation factors during blood storage entail a gradual reduction in biologic activity (6). Coagulation factor VIII and V activity experiences rapid loss during the storage period due to their lability (6). By storage of 35 days, whole blood in CPD-Adenine, factor VIII 16–20% and factor V falls to 15–21% activity. There is only slight decline observed in factor X and II during the same storage time. Other factors except for factor VII are virtually unchanged. Factor VII reduces slightly but otherwise can be activated in the cold (6). Factor VIII reduces in fresh frozen plasma that is stored for a year, and this is reduced by more rapid freezing to lower temperatures. Coagulation factors are properly maintained in platelets concentrates, with the exception of the labile factors. However, factor V falls more rapidly with room temperature storage and agitation while decline is lessened in factor VIII (6). Low coagulation factor levels can cause blood clotting to fail which leads to unexplained bleeding episodes. Healthcare practitioner can determine the cause of the bleeding and the best treatment by measuring coagulation factors (7).

This study aimed at evaluating Evaluation of changes in coagulation factors in fresh frozen plasma during storage at -18°C for 5 weeks at Kisii Teaching and Referral Hospital.. Plasma obtained from donated units eight hours post collection at KTRH is often discarded without any consideration of coagulation factor levels. Fresh frozen plasma is used without knowing the concentration of coagulation factors it contains which might cause circulatory overload without any improvement if several bags are used with low level of coagulation factors. Increased risk of venous thrombosis has been described to be associated with high plasma levels of several coagulation factors.

## Methodology

### Study site

This study was conducted at Kisii Teaching and referral hospital (KTRH) laboratory department. KTRH is located within Kisii town at the southern end of the western Kenyan highlands at an altitude of 1,660m above sea level. Coordinates for the town are 0°41’S 34°46’E / 0.683°S 34.767°E

### Sample size

The study involved 108 eligible volunteer blood donors at Kisii Satellite Blood Transfusion Center, who met the donor suitability criteria following the World Health Organization guidelines.

### Study design

This study was a longitudinal study involving time series analysis of fresh frozen plasma stored at -18°C for up to five weeks. Four hundred- and fifty-ml blood was collected into tetra blood bags containing citrate-phosphate-adenine anticoagulant-preservative (*CPDA*-*1*) as an anti-coagulant preservative for subsequent processing into fresh frozen plasma for storage at -18°C. The collected blood was centrifuged at 4000 RPM for 9 minutes within 5 - 8 hours after collection in a separate sanitized room where about 180ml plasma was formed as supernatant which then was separated and collected. The 180ml plasma obtained through centrifugation was aliquoted in three parts each containing 60ml. The first aliquot was used to assess the changes in coagulation factors in fresh frozen plasma at room temp at baseline during week one of collection (baseline), the second aliquot was used to assess the changes in coagulation factors in fresh frozen plasma storage at -18°C temp after three weeks of storage, the third aliquot was used to assess the changes in coagulation factors in fresh frozen plasma storage at -18°C temp after five weeks of storage. Coagulation factor analysis was performed using Erba Mannheim ECL 105 coagulation analyzer, India at KTRH Hematology laboratory. Thawing for subsequent coagulation factor analysis and serial testing of stored fresh frozen plasma was done using Stericox Plasma Thawing Bath, an equipment designed for rapid and uniform thawing of fresh frozen plasma (FFP) bags at 37°C, for 45 mins before the samples are analyzed by Erba Mannheim ECL 105 coagulation analyzer, India and results recorded to assess the coagulation factors changes and levels in fresh frozen plasma. Standard storage conditions for the aliquots were observed and maintained to ensure their coagulation factor levels homogeneity.

### Data management and statistical analysis

The data was recorded as numbers (value measured). Statistical analysis was descriptive statistics. The raw data collected was entered in Microsoft office Excel spreadsheet before being transferred to SPSS software version 25.0. The findings were presented in tables and graphs.

### Ethical considerations

Institutional ethical clearance was obtained from Baraton ethical review committee - (UEAB/ISERC/02/05/2022) and research permit obtained from National Commission for Science and Technology (NACOSTI) - NACOSTI/P/22/17542.

## Results

The findings were as follows;

The mean of Fresh Frozen Plasma for the first week was 114.03 with a standard deviation of 19.49. This means that most of the fresh frozen plasma factors in week one did not vary widely, but were clustered around the mean FFPW1 of the 108 blood donors: the mean FFPW3 and FFPW5 was 105.70 and 95.30 respectively as reported in Table 4.5 above. This means that there was a small and significant difference in means for the fresh frozen plasma factors for the five-week time period, hence, for the variables stated above, there was normality in distribution.

**Table 4.5:** Descriptive Statistics for Coagulation Factors in Fresh Frozen Plasma.

| Time | n | Mean | Std. Deviation | Minimum | Maximum |
| --- | --- | --- | --- | --- | --- |
| FFPW1 | 27/108 | 114.03 | 19.49 | 62.60 | 152.60 |
| FFPW3 | 87/108 | 105.70 | 19.42 | 54.80 | 144.20 |
| FFPW5 | 89/108 | 95.30 | 18.62 | 48.50 | 133.70 |

*Fresh Frozen Plasma in Week One (FFPW1), Fresh Frozen Plasma in Week three (FFPW3), Fresh Frozen Plasma in Week five (FFPW5)*

**Table 4.6:** Chi-square Test for Influence of Gender on Coagulation Factors in FFP.

|  |  |  | Gender |  | Chi-square Test |  |  |
| --- | --- | --- | --- | --- | --- | --- | --- |
|  |  |  | Male | Female | Degree of Freedom | Chi-square value | P-value |
| **W1FFP** | Normal | Count | 48 | 47 | 6 | 6.872 | 0.333 |
|  |  | % of Total | 44.4% | 43.5% |  |  |  |
|  | Abnormal | Count | 8 | 5 |  |  |  |
|  |  | % of Total | 7.4% | 4.6% |  |  |  |
| **W3FFP** | Normal | Count | 46 | 45 | 4 | 0.892 | 0.345 |
|  |  | % of Total | 42.6% | 41.7% |  |  |  |
|  | Abnormal | Count | 10 | 7 |  |  |  |
|  |  | % of Total | 9.3% | 6.5% |  |  |  |
| **W5FFP** | Normal | Count | 29 | 22 | 4 | 0.006 | 0.940 |
|  |  | % of Total | 26.9% | 20.4% |  |  |  |
|  | Abnormal | Count | 27 | 30 |  |  |  |
|  |  | % of Total | 25.0% | 27.8% |  |  |  |

**Table 4.7:**
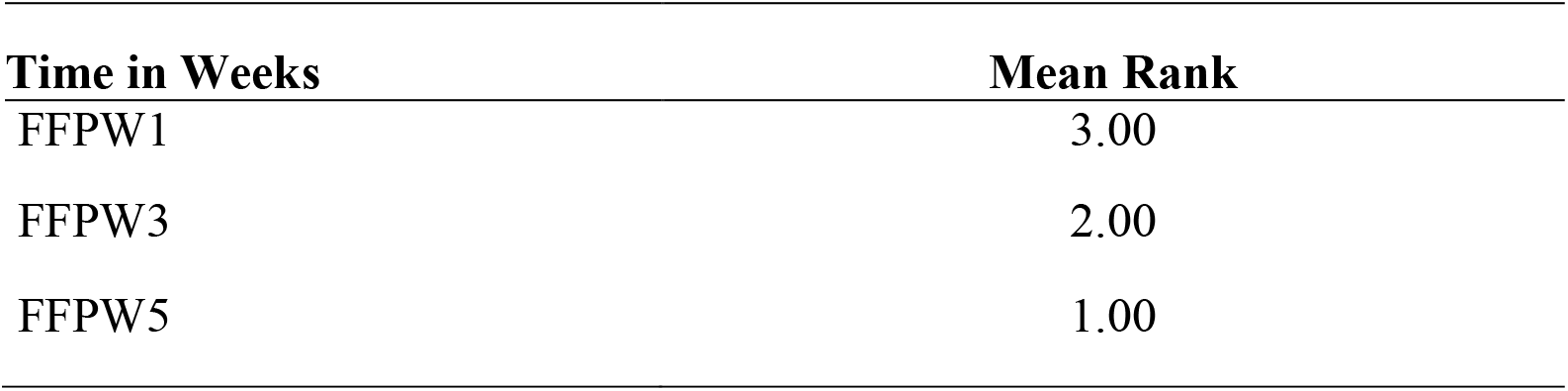
Friedman’s Test for Mean Rank.

The chi-square analysis results showed no association between gender and FFP. For the first week for the FFP for the male, 48 (44.4%) were normal and only 8 (7.4%) were abnormal, for the female, 47 (43.5%) were normal and 5 (4.6%) were abnormal. For week 5 of the study considering male, 29 (26.9%) were normal and 27 (25.0%) were abnormal and for the female, 22 (20.4%) were normal and 30 (27.8%) were abnormal. The chi-square test indicated by the p-values for three sampled weeks of the study with (0.333, 0.345 and 0.940) respectively greater than the standard alpha value of 0.05. This means that, the coagulation factors in FFP were not affected by the gender of the blood donor.

### 4.2.1 Friedman’s ANOVA Test Analysis

The Friedman test analysis was employed to establish the differences in time period in coagulation factors in fresh frozen plasma of stored blood for transfusion after blood donation by use of the mean rank test. The results were as shown below.

The mean rank for FFPW1 was 3.00, 2.00 for fresh frozen plasma for the third week after donation and 1.00 for the fifth week of storage of blood. This proves that there was a significant difference in mean ranks for the time period though in a reducing trend.

To establish the statistical significance difference between the three-selected periods of fresh frozen plasma factors, the freedman’s test statistics was applied. The results from Table 4.8 above shows that, there was a significant difference between the three selected time period for the stored blood at Kisii Teaching and Referral Hospital, Kisii County. This is shown by the chi-square value of 216.000 with asymptomatic significance value (p-value) <0.0001* less than the standard alpha 0.05.

**Table 4.8:** Friedman’s Test Statistics.

| **Test Statistics<sup>a</sup>** |  |
| --- | --- |
| **N** | **108** |
| Chi-Square | 216.000 |
| Df | 2 |
| Asymp. Sig. | .0001* |

*a. Friedman Test*

To show specific difference between the three-week timeframe for fresh frozen plasma in the donated blood, the use of Wilcoxon signed rank test was employed. The results were as shown in Table 4.9 above, which indicated a significant difference when the two variables are compared, that’s; FFPW1 and FFPW3, FFPW1 and FFW5 and lastly FFPW3 and FFPW5 all had p-value less than 0.05, thus they were significantly different from one week to another.

**Table 4.9:**
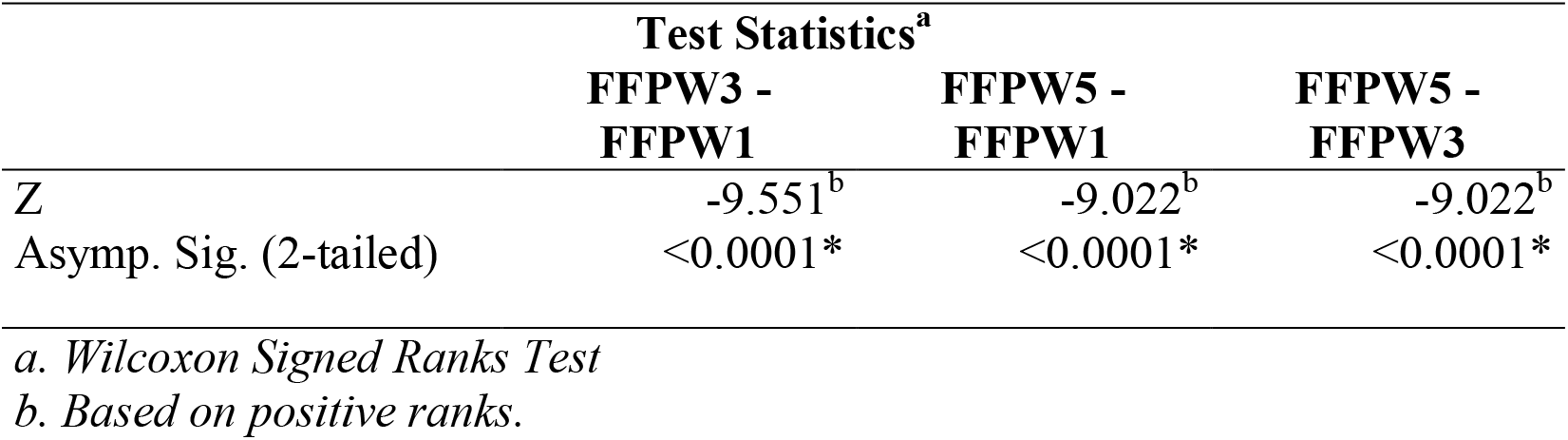
Wilcoxon Signed Ranks Test.

| **Test Statistics<sup>a</sup>** |  |  |  |
| --- | --- | --- | --- |
|  | **FFPW3 -**<br>**FFPW1** | **FFPW5 -**<br>**FFPW1** | **FFPW5 -**<br>**FFPW3** |
| Z | -9.551<sup>b</sup> | -9.022<sup>b</sup> | -9.022<sup>b</sup> |
| Asymp. Sig. (2-tailed) | <0.0001* | <0.0001* | <0.0001* |

*a. Wilcoxon Signed Ranks Test*
*b. Based on positive ranks.*

The difference in the coagulation factors in fresh frozen plasma for the donated blood in diagrammatic representation in Figure 4.7 above shows that there was a steady constant decrease in the factors as from week one with an estimated marginal mean of approximately 118 to 110 for the third week and an approximated estimated marginal mean value of 100 for the fifth week. These results are in support of the Table 4.4 results about the Friedman mean rank test which showed a constant decrease in mean ranks for the coagulation factor in fresh frozen plasma.

**Figure 4.7:**
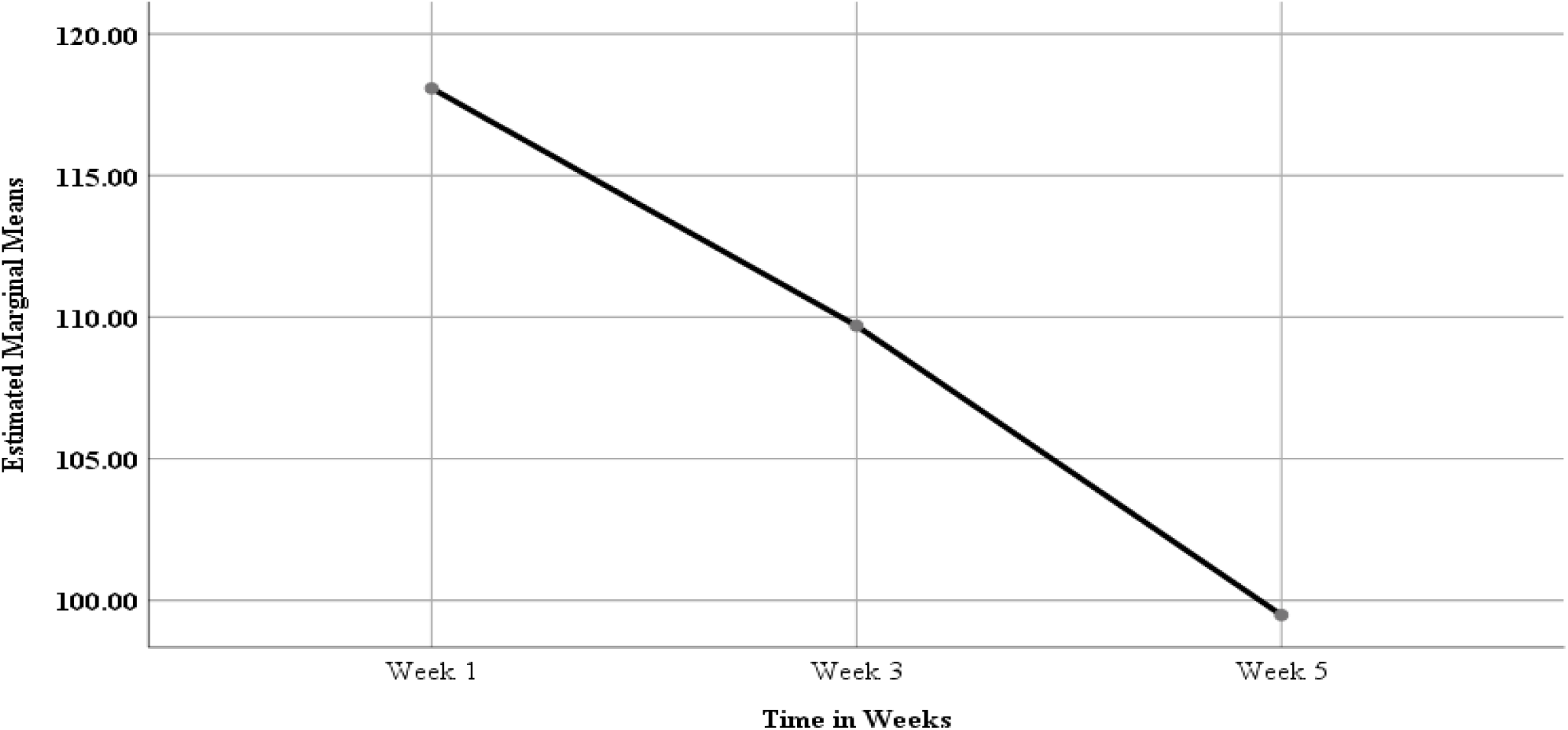
Estimated Marginal Means for Coagulation Factors in Fresh Frozen Plasma.

**Figure 4.8:**
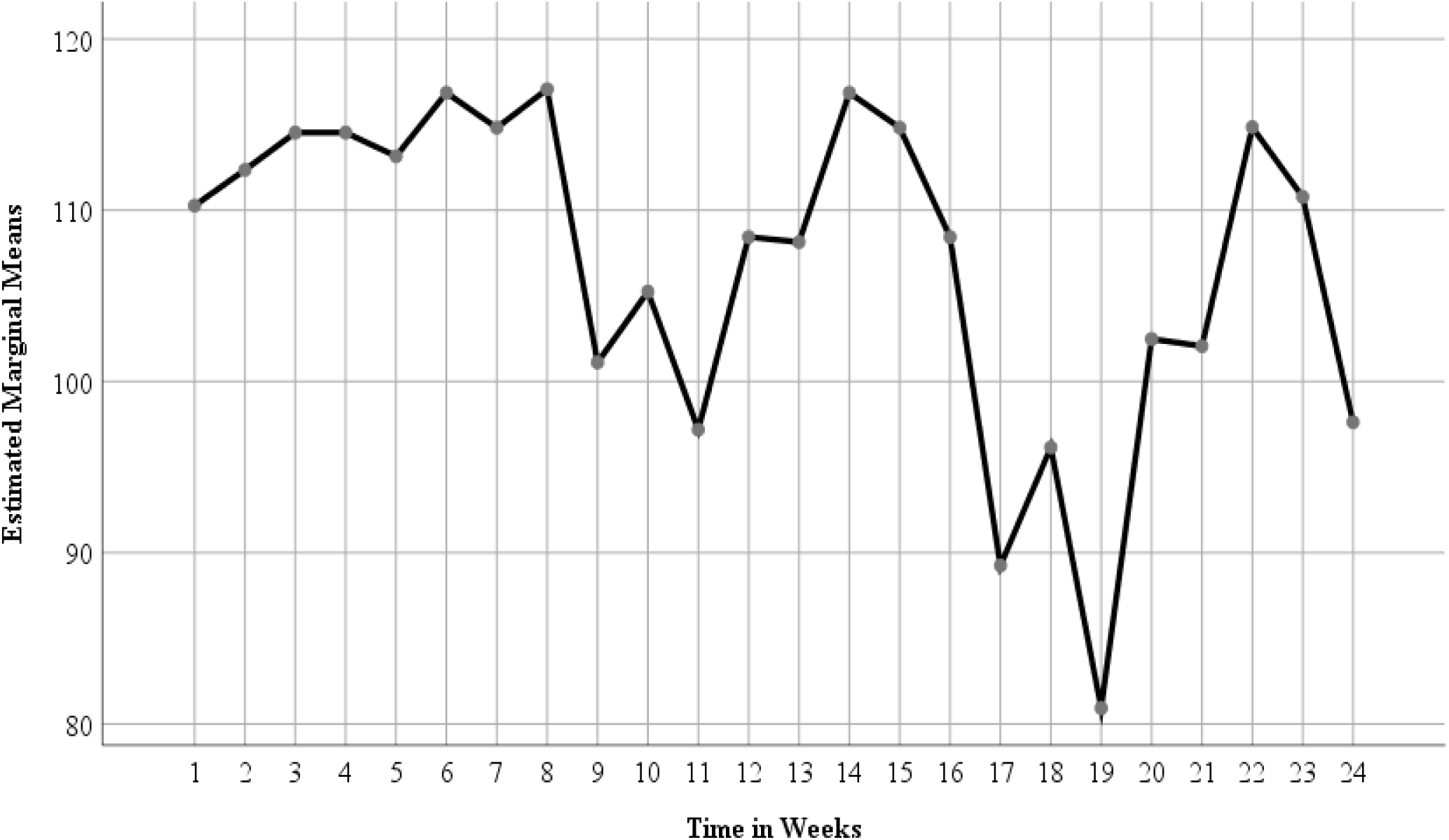
Estimated Marginal Means for Specific Coagulation Factors in Fresh Frozen Plasma.

Each week was represented by eight factors that’s; 1(Week1 FI), 2(Week1 FII), 3(Week1 FV), 4(Week1 FVII), 5(Week1 FX), 6(Week1 FXI), 7(Week1 FXII), 8(Week1 FXIII), 9(Week3 FI), 10(Week3 FII), 11(Week3 FV), 12(Week3 FVII), 13(Week3 FX), 14(Week3 FXI), 15(Week3 FXII), 16(Week3 FXIII), 17(Week5 FI), 18(Week5 FII), 19(Week5 FV), 20(Week5 FVII), 21(Week5 FX), 22(Week5 FXI), 23(Week5 FXII), 24(Week5 FXIII). This results in the line graph above of estimated marginal means and time in weeks for the coagulation factors in fresh frozen plasma shows that, at the end of the fifth week there was a decrease of coagulation factors FI with estimated marginal mean value of about 110 for the first week to the factor FXIII of week five with an estimated marginal mean value less than 100. The outcome hence means that with standardized storage conditions at the blood bank at KTRH, the coagulation factors in fresh frozen plasma were reducing though a significant decrease.

## Discussion

This objective focused on testing the change in coagulation factors in FFP from the first week of storage to the fifth week time period basing on the means and standard deviations for each timeframe. The findings showed significant changes in the coagulation factors in FFP during storage at -18 for a period to five weeks. Finding of this study were consistent with (8) who observed that that single coagulation factor activities when compared to other differed significantly hence not affecting the level of coagulation factors in the stored blood. The same results mirrored the findings of (9), that storing of blood for more than 5 day and above has the capability of reducing the coagulation factors in FFP. The study results indicated that, as the donated blood continues to be stored, there was a significant reduction in coagulation factors in fresh frozen plasma. A study done by (10), indicated that the coagulation factors decreased significantly after storage. But for better fresh frozen plasma then, thawing at 37°C has a crucial impact on the quality of the plasma content in blood before transfusion. Therefore, to avoid wastage of coagulation factors in FFP, storage of thawed plasma for a short time is advised. This is also in line with (11) that for longer storage period of blood before processing led to significant decrease in coagulation factors in FFP. The data collected analyzed suggested that there is good retention of coagulation factors in fresh frozen plasma since the different was significant. This means that, as the donated blood continues to be stored, there is a significant reduction in coagulation factors in FFP in the donated blood ready for transfusion, which also supports the results of the study with a constant reduction in mean rank from the first week to the fifth week.

## Conclusion

There was a constant decrease of coagulation factors in fresh frozen plasma during storage at - 18°C for 5 weeks at Kisii Teaching and Referral Hospital, Kisii County.

## Data Availability

All data produced in the present study are available upon reasonable request to the authors
All data analysis produced in the present work are contained in the manuscript

## Data availability

All data are shared on this manuscript

## References

1. Craik, C. S., Page, M. J., & Madison, E. L. (2011). Proteases as therapeutics. Biochemical Journal, 435(1), 1–16.

2. Storch, E. K., Custer, B. S., Jacobs, M. R., Menitove, J. E., & Mintz, P. D. (2019). Review of current transfusion therapy and blood banking practices. Blood Reviews, 38, 100593.

3. Rinchard, J., Dabrowski, K., Garcia–Abiado, M. A., & Ottobre, J. (1999). Uptake and depletion of plasma 17α-methyltestosterone during induction of masculinization in muskellunge, Esox masquinongy:: Effect on plasma steroids and sex reversal. Steroids, 64(8), 518–525.

4. Lee, J.-H., Kim, S.-K., Khawar, I. A., Jeong, S.-Y., Chung, S., & Kuh, H.-J. (2018). Microfluidic co-culture of pancreatic tumor spheroids with stellate cells as a novel 3D model for investigation of stroma-mediated cell motility and drug resistance. Journal of Experimental &Clinical Cancer Research, 37(1), 1–12.

5. British Committee for Standards in Haematology Chairman), B. T. T. F. (J. D., O’Shaughnessy, D. F., Atterbury, C., Bolton Maggs, P., Murphy, M., Thomas, D., Yates, S., & Williamson, L. M. (2004). Guidelines for the use of fresh-frozen plasma, cryoprecipitate and cryosupernatant. British Journal of Haematology, 126(1), 11–28.

6. Simon, T. L. (1988). Changes in plasma coagulation factors during blood storage. Plasma Therapy and Transfusion Technology, 9(3), 309–315.

7. Dahlbäck, B. (2000). Blood coagulation. The Lancet, 355(9215), 1627–1632.

8. Gratz, J., Ponschab, M., Iapichino, G. E., Schlimp, C. J., Cadamuro, J., Grottke, O., Zipperle, J., Oberladstätter, D., Gabriel, C., & Ziegler, B. (2020). Comparison of fresh frozen plasma vs. coagulation factor concentrates for reconstitution of blood: An in vitro study. European Journal of Anaesthesiology| EJA, 37(10), 879–888.

9. Noordin, S. S., Karim, F. A., Mohammad, W. M. Z. bin W., & Hussein, A. R. (2018). Coagulation factor activities changes over 5 days in thawed fresh frozen plasma stored at different initial storage temperatures. Indian Journal of Hematology and Blood Transfusion, 34(3), 510–516.

10. Kuta, P., Melling, N., Zimmermann, R., Achenbach, S., Eckstein, R., & Strobel, J. (2019). Clotting factor activity in fresh frozen plasma after thawing with a new radio wave thawing device. Transfusion, 59(5), 1857–1861.

11. Dogra, M., Sidhu, M., Vasudev, R., & Dogra, A. (2015). Comparative analysis of activity of coagulation Factors V and VIII and level of fibrinogen in fresh frozen plasma and frozen plasma. Asian Journal of Transfusion Science, 9(1), 6.

